# Measuring the accuracy of electronic health record (EHR)-based phenotyping in the *All of Us* Research Program to optimize statistical power for genetic association testing

**DOI:** 10.1101/2025.07.30.25332455

**Authors:** John Baierl, Yi-Wen Hsiao, Michelle Jones, Pei-Chen Peng, Paul D. P. Pharoah

## Abstract

Accurate phenotyping is an essential task for researchers utilizing electronic health record (EHR)-linked biobank programs like the *All of Us* Research Program (*AoU*) to study human genetics. While their large cohort sizes offer increased statistical power for detecting novel risk alleles, those benefits are undermined if participants’ disease status cannot be accurately determined from EHRs. Little guidance is available on how to select an EHR-based phenotyping procedure that maximizes downstream statistical power. We used observed carrier frequencies of known risk genes for ovarian, female breast, and colorectal cancers to estimate the accuracy of EHR-based phenotyping strategies for each disease in *AoU* (v7). We found that the choice of phenotype definition can have a substantial impact on statistical power for association testing, particularly for rarer diseases. Additionally, our results suggest that the accuracy of higher-complexity phenotyping algorithms is inconsistent across Black and non-Hispanic White participants in *AoU*, highlighting the potential for case ascertainment biases to impact downstream association testing. We discuss the implications of this as well as potential mitigation strategies.

## Introduction

Large-scale biobank programs like UK Biobank and the *All of Us* Research Program (*AoU*) play a central role in the study of human genetics and disease prediction by providing researchers with low-cost access to hundreds of thousands of whole genomes. These large cohort sizes offer the potential to uncover novel genetic associations undetectable in smaller study designs. This is critical for detecting novel risk alleles of modest effect size or those that are rare in the population. However, these data often rely on the health record databases made available to researchers for accurate identification of phenotypes. For example, researchers in *AoU* rely primarily on electronic heath records (EHRs) to infer disease status.

This poses many challenges. EHRs are known to contain inaccuracies and are often incomplete with record-keeping norms that vary by hospital system and location, making it difficult to determine the optimal criteria for reliably identifying cases^1–3^. Many possible approaches to EHR-based case ascertainment exist. The simplest method is to select all individuals with any observation of a qualifying list of diagnosis codes. The Cohort Builder tool provided on the *AoU* Researcher Workbench implements this strategy. Phenotyping based on a minimum of one instance of a qualifying diagnosis code in the EHR is common in practice^4–8^. Adopting this phenotype definition can be expected to maximize sensitivity for detecting cases by including all individuals with any disease record in the database as cases. However, given the known inaccuracies of EHR data and the use of rule out codes in ICD-9-CM and ICD-10-CM, such an admissive criterion risks including false positives in the case set, potentially sacrificing downstream statistical power due to lowered phenotyping specificity^9,10^.

More sophisticated EHR-based methods seek to improve phenotyping specificity by imposing stricter requirements on the strength of evidence needed to qualify as a case. One approach is to apply a more complex algorithm-based EHR phenotyping procedure to reduce the likelihood of false positives. Many such phenotyping pipelines have made publicly available to researchers, such as those developed through the Electronic Medical Records and Genomics (eMERGE) Network^11^. Another approach is to require the occurrence of multiple qualifying diagnosis codes on distinct days to qualify as a case, or the so-called “rule of two”^12,13^.

While these additional requirements may limit false positives in the final case set, they also run the risk of decreasing statistical power by reducing sample sizes if they are overly conservative and exclude many true positives. Little guidance is available about the degree to which different approaches tradeoff sensitivity for specificity, how disease-specific these tradeoffs are, and the implications for maximizing power for downstream association analysis. While the gold standard for evaluating phenotyping accuracy is manual chart review, such an undertaking is impossible in many large biobanks like *AoU*, forcing researchers using these platforms to rely on other metrics or heuristics.

In this paper, we evaluate case sets in *AoU* for epithelial ovarian cancer (EOC), female breast cancer, and colorectal cancer (CRC) developed using three different criteria. We use observed carrier frequencies of pathogenic germline mutations in known high-penetrance risk alleles to estimate the misclassification rates under the more sensitive but less specific case definitions, then compare downstream statistical power to the smaller, higher-specificity case set for each disease. While this analysis is most directly applicable to users of *AoU*, the general concerns and methodology in assessing and selecting an optimal phenotype definition are broadly relevant to researchers utilizing biobank data and EHR databases.

## Materials and methods

### Phenotyping algorithms

We compared three approaches to EHR-based phenotype ascertainment from the *AoU* short-read whole genome sequencing (srWGS) cohort: (1) identifying case status based on at least one instance of a qualifying diagnosis code (single EHR); (2) identifying any participant with two occurrences of a qualifying code on different calendar days as a case (rule of two), and (3) utilizing a more complex algorithm published through the eMERGE network. The rule of two comes from the PheWAS (phenome-wide association study) literature targeting a consistent and easy-to-implement criteria for defining cases across a wide range of diseases^12,13^. Requiring a minimum of two qualifying diagnosis codes (Phecodes) has been found to reasonably balance sensitivity and specificity^14^.

Algorithm-based phenotyping methods employ a more complex logical structure that varies by target disease and may require more diverse EHR evidence such as drug or procedure records in addition to diagnosis codes^15,16^. We tested the validated phenotyping algorithms for EOC, breast cancer, and CRC published through the eMERGE network. These algorithms are commonly used in practice and serve as a useful benchmark, with several implementations published directly on the *AoU* researcher workbench for use on the platform. Full lists of qualifying codes and further detail on pipeline structure, reasoning, and validation are available through PheKB^17–19^. Details on specific implementation in the *AoU* data model are available in the Supplementary Materials. Algorithm logic for identifying ovarian, breast, and colorectal cancers is pictured in *fig, 1*. The breast and ovarian cancer pipelines rely exclusively on condition and observation code occurrences which are available in the *AoU* EHR database. Though the published CRC algorithm can incorporate cancer registry data that is unavailable in *AoU*, the authors specify that this is not required for its implementation to assemble a case set^19^.

**Figure 1.**
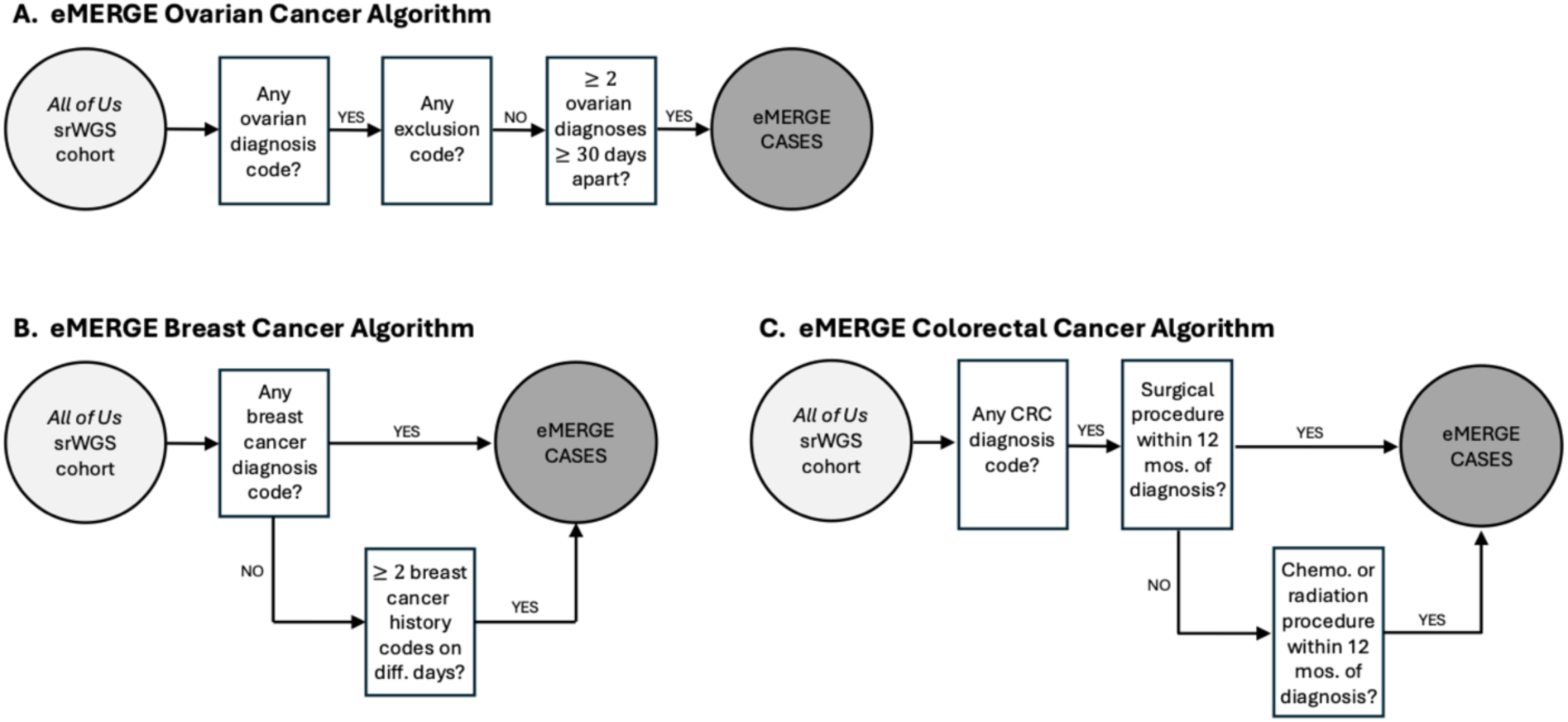
Algorithm-based eMERGE phenotyping logic. Algorithm logic for the three eMERGE phenotypes in this analysis: (**A**) ovarian cancer, (**B**) breast cancer, and (**C**) colorectal cancer. Individuals with some qualifying diagnosis code but otherwise falling short of the eMERGE criteria were considered tentative cases under the single EHR criteria. Control set logic and optional steps incorporating cancer registry data (unavailable in AoU) are not shown.

Non-cancer control sets were constructed from the set of participants with srWGS available who had consented to link their EHRs with their genotyping data and had no record of any cancer-related diagnosis code (“malignant neoplastic disease” or any descendant concept). This set was further filtered for each phenotype based on a list of control exclusion codes provided in each disease-specific algorithm. While pathology reports were unavailable to screen controls, our conservative filtering of controls based on any instance of a cancer-related code in the EHRs is likely to eliminate most false negatives.

Our goal is to assess which case definition maximizes downstream statistical power for genetic association study in *AoU*. For each disease, we compare a high-specificity phenotyping approach with the two others as candidate higher sensitivity alternatives. This produces four different groups of individuals for each phenotype (*fig. 2*): non-cancer controls (*n*_0_), confident cases (*n*_1_), and two sets of tentative cases 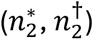 consisting of individuals who met the candidate alternative case criteria but fell short of qualifying under the high-specificity definition. Statistical power under each case definition depends on its probability of admitting false positives. To estimate the number of affected individuals in each tentative case set, we leverage the observed carrier frequencies of known high-penetrance risk alleles.

**Figure 2.**
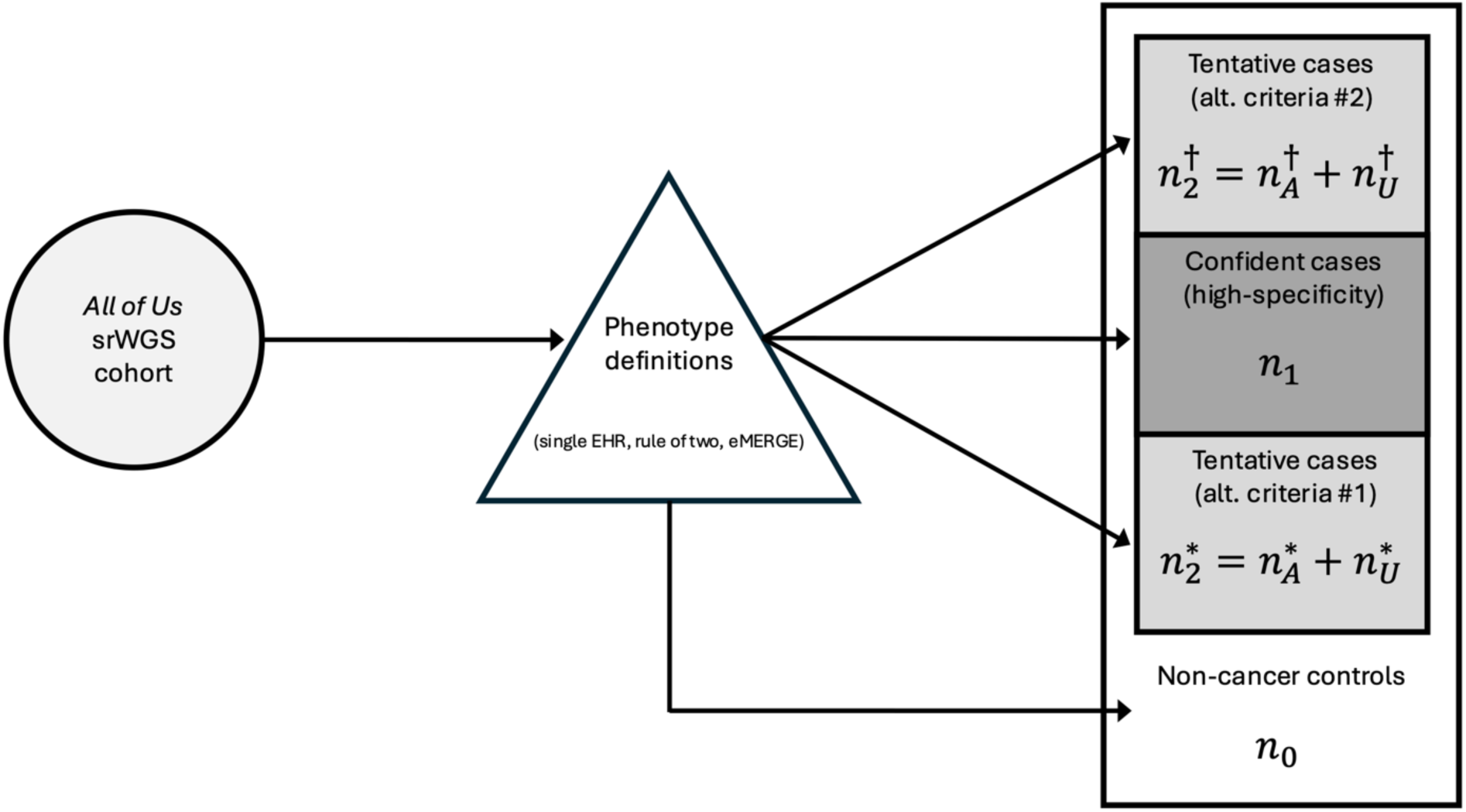
Data processing overview and simplified Venn diagram with case set notation. For each phenotype, three different case sets are produced by the single EHR, rule of two, and eMERGE definitions. The tentative case sets consist of individuals who qualified under the higher-specificity definition (varies by phenotype), but not the more sensitive alternative. These groups consist of an unknown mixture of affected (*n*_*A*_) and unaffected (*n*_*U*_) individuals. Note that the two sets of tentative cases are not mutually exclusive since some participants may qualify as cases under both definitions.

### Estimating misclassification rates and statistical power

Ovarian, breast, and colorectal cancers all have known genes with high-penetrance risk alleles that confer substantial lifetime risks for each disease. While pathogenic variants in these genes are highly informative for predicting an individual’s lifetime disease risk, their observed frequencies in the population and in unselected case series can also serve as a benchmark for identifying the degree to which a set of tentative cases is expected to contain truly affected individuals. For example, *BRCA1* and *BRCA2* (*BRCA1/2*) have been extensively studied as susceptibility genes for ovarian cancer, conferring average cumulative risk by age 70 of 39% and 11% respectively^20^. While rare in the general population outside of subpopulation-specific founder variants (e.g. Ashkenazi Jewish populations), pathogenic *BRCA1*/*2* variants are much more common among EOC cases^21–23^. Population-based studies find that 5-15% of EOC cases are expected to carry either a predicted loss of function (pLoF) *BRCA1*/*2* variant relative to 0.2% in the general population^24,25^.

Discrepancies between the observed frequencies of pathogenic *BRCA1/2* variants in a tentative case group and a set of truly affected individuals contains information about the degree to which that group contains true positive cases. Let *f0*, *f1*, and *f2* denote the observed pooled predicted loss of function carrier frequencies in non-cancer controls, true positive cases, and tentative cases, respectively. By expressing *f2* as a weighted average of the frequencies in true cases and controls, the expected number of affected individuals in the tentative group 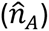 can be estimated by:

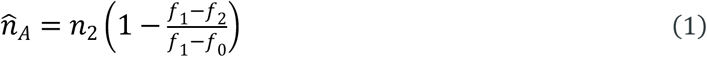

From this, desired attributes of the tentative case set such as the number of unaffected individuals 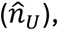 specificity, positive predictive value (PPV), or the probability of a randomly selected case being unaffected are straightforward to compute. We assume perfect specificity in the confident case set, taking *f1* to be an estimate of the carrier frequency in affected individuals and *f0* to be an estimate of the carrier frequency in unaffected individuals. While the latter assumes that none of the non-cancer controls are truly affected, our conservative exclusion criterion makes violations of this assumption likely to be small.

Note that this estimator does not account for differences in genetic ancestry between cases and controls. However, outside of subpopulation-specific founder variants (e.g. *BRCA1* in Ashkenazy Jewish populations), the variation across the ancestry continuum in carrier frequencies of the high-penetrance risk alleles considered here is generally small relative to the frequency differences between cases and controls^26,27^. Thus, population structure is likely to have a limited impact on the utility of this estimate.

The high-risk genes used to estimate phenotyping accuracy for each disease are presented in *table 1*, along with their corresponding lifetime average risks and estimated frequencies in true positive cases. Like ovarian cancer, *BRCA1*/*2* mutations confer substantial lifetime risk of breast cancer. Mismatch repair (MMR) deficiency or Lynch Syndrome is highly penetrant for CRC. Approximately 3% of CRC cases are expected to carry pathogenic germline variants in a MMR gene (*MLH1*, *MSH2*, and *MSHC*)^28^. We applied a standard definition of protein-truncating mutations, considering nonsense variants, frameshift indels, variants predicted to disrupt the consensus splice site, and pathogenic missense variants to be pLoF. Missense variant pathogenicity was based on ClinVar classification; variants listed as either “pathogenic” or “likely pathogenic” with no conflicts among reviewers were classified as pLoF.

**Table 1.** Risk genes used for phenotyping assessment.

| Phenotype | Risk gene | Lifetime avg. risk | Frequency in cases |
| --- | --- | --- | --- |
| Ovarian cancer | *BRCA1* | 39%<sup>20</sup> | 5-15%<sup>25</sup> |
|  | *BRCA2* | 11%<sup>20</sup> |  |
| Breast cancer | *BRCA1* | 65%<sup>20</sup> | 5-10%<sup>29</sup> |
|  | *BRCA2* | 45%<sup>20</sup> |  |
| Colorectal cancer | *MLH1* | 41%<sup>30</sup> | 3%<sup>28</sup> |
|  | *MSH2* | 48%<sup>30</sup> |  |
|  | *MSH6* | 12%<sup>30</sup> |  |

Calculating statistical power to detect risk alleles under outcome misclassification has been previously studied^31^. We apply this approach to the loss of function burden test setting, where a logistic regression model is fit to test association between binary loss of function status in the target gene with the disease of interest (see Supplementary Materials for details).

## Results

### Case set characteristics and phenotyping accuracy

Case set sizes from implementing the three phenotype definitions in *AoU* are presented in *table 2*. Demographic characteristics stratified by disease and case status are shown in *table S1* and *fig. S2*. The eMERGE algorithms were generally the most complex and restrictive across the target diseases, requiring the greatest depth and variety of EHR evidence to be classified as a case. For example, in addition to requiring multiple occurrences of a qualifying diagnosis code, the eMERGE EOC algorithm also restricts cases to diagnoses separated by at least 30 calendar days. Breast cancer was an exception, with the rule of two definition producing the highest level of specificity. We consider the eMERGE definition to produce the confident case sets for EOC and CRC, and the rule of two to produce the confident case set for breast cancer. The remaining phenotyping approaches serve as the two tentative case sets for each disease.

**Table 2.** Counts and pooled pLoF carrier frequencies for confident case, tentative case, and control sets in AoU v7. Score-based S1% confidence bounds for frequencies are provided. Note that the confident cases are a subset of the complete case sets under the higher sensitivity definitions, and that the two tentative case sets are not mutually exclusive.

|  | Confident cases |  | Tentative cases: set 1 |  | Tentative cases: set 2 |  | Controls |  |
| --- | --- | --- | --- | --- | --- | --- | --- | --- |
|  | Count | Freq. | Count | Freq. | Count | Freq. | Count | Freq. |
| Ovarian cancer | 368 | 9.5%<br>[7.2%, 12.4%] | 175 | 6.9%<br>[4.3%, 10.8%] | 513 | 4.5%<br>[3.2%, 6.3%] | 95,342 | 0.5%<br>[0.5%, 0.6%] |
| Breast cancer | 5,501 | 4.0%<br>[3.5%, 4.4%] | 899 | 3.6%<br>[2.7%, 4.8%] | 946 | 3.5%<br>[2.6%, 5.5%] | 95,575 | 0.5%<br>[0.5%, 0.5%] |
| Colorectal cancer | 209 | 1.9%<br>[0.8%, 4.3%] | 896 | 2.9%<br>[2.1%, 4.0%] | 1,644 | 1.9%<br>[1.4%, 2.5%] | 157,802 | 0.1%<br>[0.1%, 0.2%] |

Phenotype definition: = eMERGE = Rule of two = Single EHR

There were considerable differences in case set sizes across methods and target disease. The smallest differences occurred in breast cancer phenotyping, with the eMERGE and single EHR criteria returning very similar numbers of cases. The rule of two returned 15% fewer breast cancer cases than the eMERGE algorithm, which imposes no multiple observation requirement for diagnosis codes. For both EOC and CRC, the eMERGE algorithm was the most restrictive, returning the fewest number of cases with the rule of two case sets falling between the eMERGE and the single EHR criteria. CRC phenotyping provided the greatest contrast between methods. Distributions in age at diagnosis (*fig. 2*) showed little difference between case definitions across ovarian, breast, and colorectal cancer cases.

The substantial disparity in CRC case counts between definitions (*table 2*) highlights the challenges in developing and validating a single algorithm to apply to EHR databases spanning a range of health care systems with varying degrees of missingness. A validated case definition optimized to reasonably balance sensitivity and specificity in one setting may perform very differently in another. The rule of two effectively serves as a middle ground here, splitting the difference between the single EHR and eMERGE approaches.

Pooled pLoF carrier frequencies by case status and phenotype are presented in *table 2*. For EOC and breast cancer, the confident case sets showed the highest carrier frequencies of the three tested case sets, with the observed values for both diseases aligning with population-based estimates in true cases (*table 1*). For EOC, frequencies in both the rule of two and single EHR tentative case sets are lower than in the confident cases but considerably higher than the frequency in non-cancer controls, suggesting that both groups likely contain a mixture of truly affected and unaffected individuals. Notably, carrier frequencies are lower in the single EHR group, which aligns with claims that relying on a single diagnosis code risks admitting unaffected individuals and that requiring a more rigorous degree of EHR-based evidence is likely to reduce false positives. Passing these empirical frequencies to *eq. 1*, we estimate that 226 women are truly affected with EOC (91%-CI: [132, 359]) among the 513 women in the single EHR tentative case set, and that 123 women are truly affected (91%-CI: [60, 123]) among the 175 in the rule of two tentative case set.

For breast cancer, there was little difference in pooled *BRCA1/2* carrier frequencies between the set of confident cases from the rule of two and the tentative cases from the single EHR criteria. Note that there was considerable overlap between the tentative breast cancer sets, with only 47 participants qualifying under the single EHR criteria that did not qualify under eMERGE (*fig. S3*), so we primarily focus on the single EHR set as the alternative to the rule of two in subsequent breast cancer analysis for simplicity. By *eq. 1*, we estimate a PPV of 98% (91%-CI: [94%, 100%]) under the single EHR criteria. Of these 946 single EHR cases, 927 of these women are expected to be truly affected with breast cancer (91%-CI: [885, 946]).

Carrier frequencies of pLoF MMR mutations in CRC cases suggest that the eMERGE case definition is likely overly conservative in *AoU*. 1.9% (91%-CI: [0.8%, 4.3%]) of CRC eMERGE cases carry a pLoF variant in any MMR gene (*table 2*). The prevalence of Lynch Syndrome in CRC cases is estimated between 2% and 3% in the general population, so this result again aligns with expectation in true cases^32^. Both tentative case sets show comparable or greater carrier frequencies than the eMERGE set, with 3.2% of rule of two tentative cases (91%-CI: [2.4%, 4.4%]) and 2.1% of single EHR cases (91%-CI: [1.6%, 2.8%]) carrying a pathogenic MMR variant. While the frequency is higher in the rule of two tentative case set than in the eMERGE case set, there is considerable uncertainty in the latter estimate due to the small sample size. There is no evidence of a difference in frequencies between these two case sets (*P* = 0.43). Overall, results show little evidence of misclassification in the tentative CRC case sets. Estimates from *eq. 1* predicts perfect classification accuracy for both the rule of two (PPV = 100%; 91%-CI: [52%, 100%]) and single EHR (PPV = 100%; 91%-CI: [83%, 100%]) definitions as a result.

## Power analysis

*Fig. 3* compares the power to detect novel risk alleles at the *α* = 5.0 × 10^−4^ significance level in *AoU* v7 at these estimated false positive proportions in the two higher-sensitivity case sets with the confident case set. *Fig. 3* also depicts a hypothetical optimal classification scheme that ascertains all the cases in the confident set as well as the estimated true cases in the tentative set with perfect specificity. This estimated upper bound depicts how much power is being lost by subpar case identification among the tentative cases. Three known risk genes for EOC are plotted in *fig. 3* for reference.

**Figure 3.**
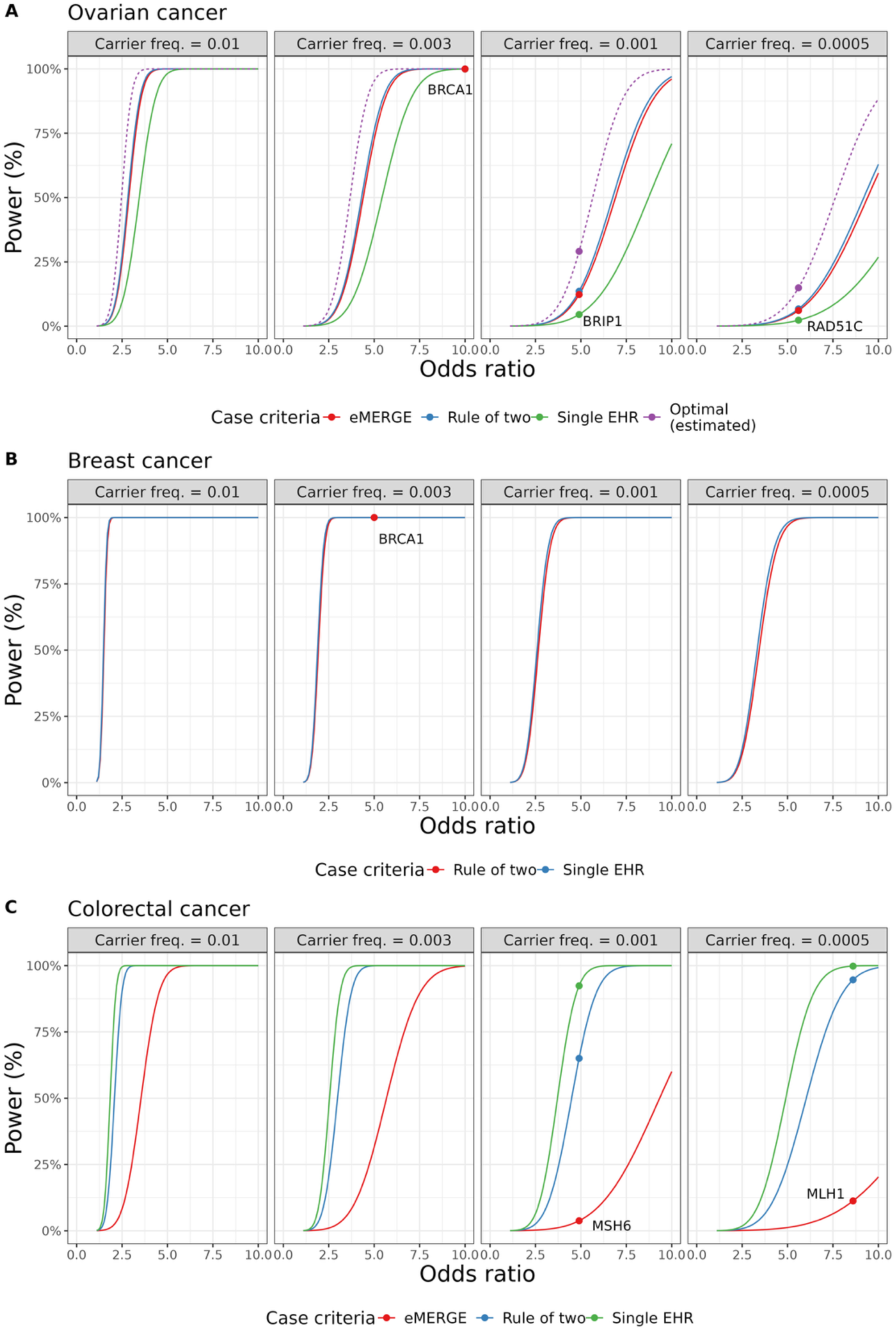
Power to detect risk alleles at *α* = 5.0 × 10^−4^ significance level by phenotype and target variant frequency and effect size (odds ratio). Note that the eMERGE case set is ommitted from the breast cancer plots (**B**) due to the large overlap with the single EHR case set. The estimated optimal classification is omitted from breast cancer (**B**) and colorectal cancer (**C**) plots since the single EHR case set is predicted to achieve a high degree of specificity for those diseases.

We estimate that the eMERGE and rule of two case definitions yield comparable statistical power for EOC association analysis, with a marginal improvement in power under the rule of two. The single-EHR case criterion reduces power due to the higher number of false positives in that case set. Though the rule of two identifies additional cases with reasonably good accuracy, the predicted number of false positives is sufficient to offset the gain in power from increasing the case set size, resulting in comparable power to the eMERGE algorithm.

While the loss of power under the single EHR definition is generally small for common variants, this difference can be substantial for rare variants. For example, researchers relying on the single EHR case criteria would see a loss in power of 37% relative to the rule of two (30% versus 67%) for detecting a novel rare risk allele with the frequency of *BRIP1* (*f* = 0.001) and an odds ratio (OR) of 7.5. Selecting an optimal phenotyping approach could be a make-or-break choice for study design in this setting. Notably, all three phenotyping approaches here are predicted to achieve power substantially below the estimated optimal phenotyping in *AoU*, suggesting that imperfect phenotyping is likely limiting the power of the *AoU* cohort for rare variant association analysis in EOC cases and controls. For example, an association analysis would achieve an estimated 91% power under optimal phenotyping for the same hypothetical variant above (*OR* = 7.5, *f* = 0.001).

For breast cancer analysis, the difference in power between phenotyping methods was much smaller than for EOC. The eMERGE case criteria is omitted from *fig. 3* due to redundancy with the single EHR breast cancer definition. We predict that the single EHR definition achieves a modest in improvement in power resulting from a 17% increase in case set size while admitting a small number of predicted false positives. However, the choice of case definition is unlikely to be consequential for analysis in *AoU*.

Similarly, differences in power for CRC analysis were entirely due to differences in sample size because of the estimated perfect classification under the tentative phenotype definitions. However, the differences in case set sizes for CRC were considerable. Power was lowest under the eMERGE case definition, and relying on this phenotyping approach in *AoU* is likely to result in an underpowered study. For example, an analysis relying on the eMERGE CRC definition would have power to detect the known association with *MLH1* of only 11% compared to 95% under the rule of two (91%-CI: [89%, 95%]) and 99.8% under the single EHR definition (91%-CI: [95%, 99.9%]). The single EHR definition results in the greatest power due to the large case set size while retaining a high degree of specificity. The difference between the rule of two and single EHR criteria is considerably smaller than the difference with eMERGE.

As the *AoU* cohort continues to grow toward its stated goal of one million participants, power to detect novel risk alleles will be improved. To assess the degree to which the concerns discussed here are likely to remain relevant even at this large cohort size, we project these estimates from *AoU* v7 (∼245,000 participants with whole genome sequencing) to a cohort size of one million. When scaling these calculations, we hold disease prevalence and classification accuracy of the phenotype definitions constant.

Results are shown in *fig. 4*. The differences in power between phenotype definitions remain substantial even at these large study designs. This is particularly acute for rare and very rare risk alleles of moderate penetrance. For example, power to detect an association with the known EOC risk gene *RAD51C* is 38% lower under the single-EHR criteria than the rule of two (32% versus 70%) and far below the estimated power under optimal phenotyping (93%). Even at this large cohort size, all three phenotyping methods are projected to produce an underpowered study to replicate a known EOC risk gene relative to what may be achievable under optimal phenotyping in *AoU*.

**Figure 4.**
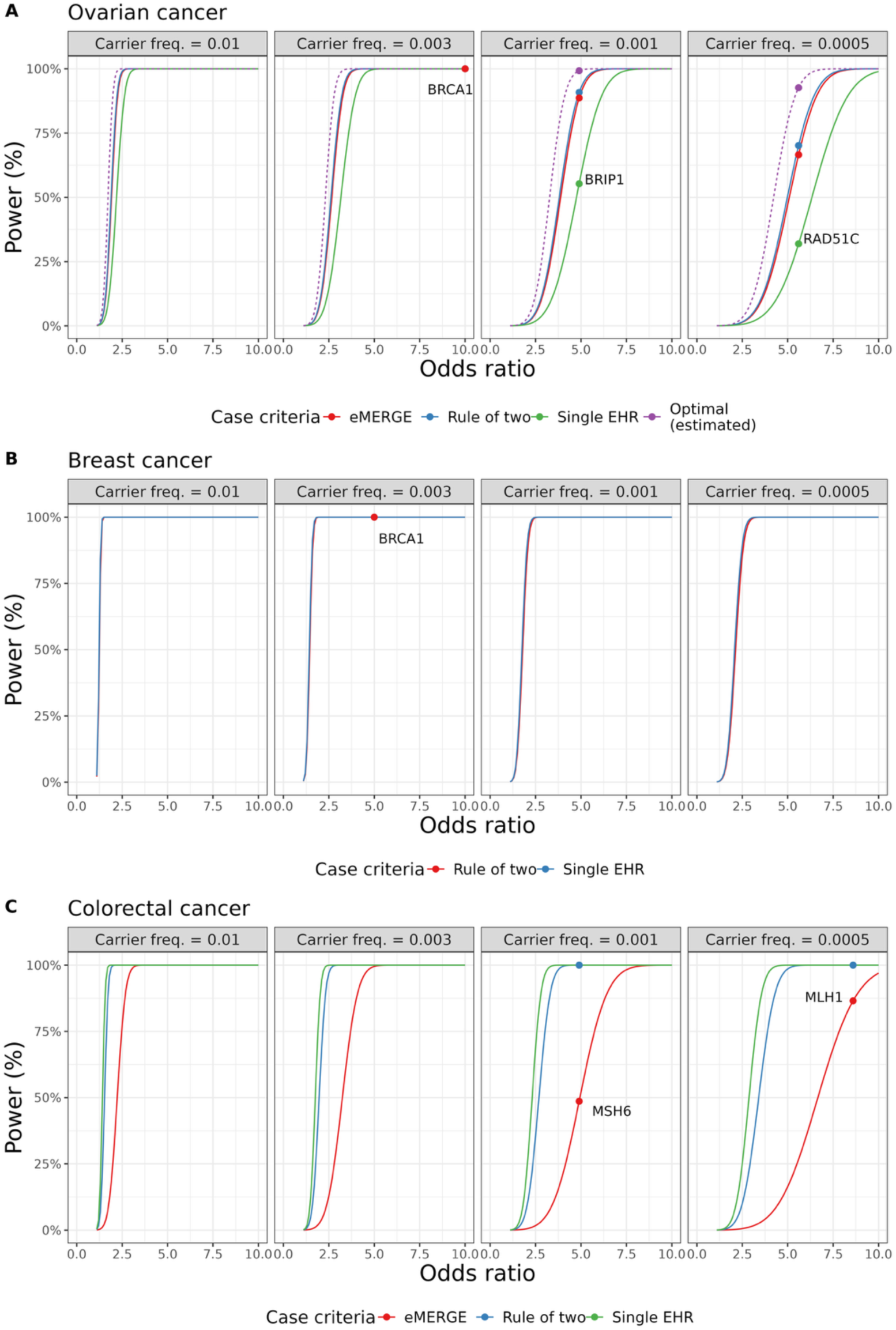
Power to detect risk alleles at *α* = 5.0 × 10^−4^ significance level by phenotype and target variant frequency and effect size (odds ratio) after projecting AoU cohort to 1 million participants. The cohort projection holds the disease prevelance and classification accuracy of each case definition constant, scaling up the case set size.

## Discussion

In this study, we leveraged observed carrier frequencies of known risk genes for ovarian, female breast, and colorectal cancers to estimate the accuracy of EHR-based phenotyping in *AoU* and select case sets most likely to optimize downstream statistical power. These results highlight how disease prevalence, EHR phenotyping accuracy, and target variant characteristics interact to determine the optimal EHR-based case definition for genetic association analyses. There was no one-size-fits-all best approach predicted to maximize statistical power across all phenotypes in *AoU*. The differences were most pronounced for the rarer diseases (EOC, CRC) while all methods achieved comparable power for breast cancer association analysis. One reason for this is predicted by theory. The cost to statistical power of including false positives in the case set increases sharply with decreasing disease prevelance^31^. This suggests that for rare diseases, more emphasis should be given to maximizing specificity when selecting a phenotype definition to minimize false positives. However, the need to maximize study size for studying rare phenotypes makes large biobanks like *AoU* appealing. This tension complicates selecting a phenotype definition in this setting. The procedure outlined here serves as a useful benchmark for informing such decisions.

EHR practices and reliability are heterogeneous across health care systems, so the best practices for phenotype ascertainment are likely to be both disease- and cohort-specific. For instance, UK Biobank links genomic data with a national cancer and pathology reports, enabling more reliable case identification. However, the concerns discussed here are broadly relevant across cohort studies that rely on patient EHRs or self-reported surveys. Phenotyping decisions must weigh factors like disease prevalence, depth of EHR-based evidence required by the case definition, and the completeness of the specific EHR database. For example, while the eMERGE CRC algorithm was found to be overly conservative in *AoU*, it may be well-suited for contexts with more complete EHRs of chemotherapy and surgical procedures.

The proportions of non-Hispanic White (referred to hereafter as White) and Black individuals in each case set differed substantially by case definition (*table S1*). More stringent phenotype definitions tended to bias case sets to be disproportionately White relative to non-cancer controls across all diseases. This is consistent with the EHR literature finding that incompleteness and biased data collection in EHRs mirrors structural inequalities^2,3^. The observed discrepancies in *AoU* are likely artifacts of this systemic bias in EHRs rather than true differences in disease risk by race or genetic ancestry. This implies that demographic characteristics of the cohort are likely to impact the performance of phenotype definitions by shaping the degree of completeness in the EHR database. In the context of the stated objective of *AoU* to diversify participation in biomedical data, this disparity is particularly concerning^33^. Efforts to enhance diversity in genetic research are undermined if phenotyping accuracy is biased toward White participants, potentially limiting the discovery of novel risk alleles more common in non-EUR-origin populations.

Our power calculations assume independence between phenotyping errors and target genotypes. While this is a reasonable assumption for the study of variants that do not show large frequency differences across the ancestry continuum, it is likely not the case for all applications. This poses a challenge for researchers performing association testing with predictors that vary by genetic ancestry. Polygenic scores (PGSs) are one example. PGS values can vary considerably by ancestry^34,35^. A PGS association analysis relying on a high-specificity phenotyping algorithm that systematically gives more sensitive disease identification for EUR-origin populations would be subject to differential misclassification in which the outcome errors are dependent on the predictor of interest. While non-differential misclassification is expected to bias effect size estimates toward the null, this is not true in the differential misclassification setting^36,37^. Researchers should be aware for the potential of EHR-based case ascertainment to induce such a bias in their analyses, and that this is likely to increase under more complex phenotyping procedures.

Facilitating accurate phenotyping of rare disease should be a point of emphasis for *AoU* moving forward to allow the full benefits of the size and diversity of this cohort to be realized. Since relying on EHRs carries inherent limitations, supplementing with orthogonal information may be useful to grow case set sizes in settings where maximizing power is critical. Self-reported disease status through survey data may serve as a useful supplement to EHR data.

*AoU* administers an optional Personal and Family Health History survey where participants can indicate whether they or a family member have received a previous diagnosis of a wide range of diseases. 47% of *AoU* srWGS participants completed this survey. *Table 3* shows counts and frequencies for the same case and control sets as *table 2* but further stratified by whether they indicated a prior diagnosis in a survey response. While the accuracy of self-reported disease status is difficult to validate, the elevated frequencies of high-risk alleles in the tentative case sets among surveyed individuals reporting a prior diagnosis suggests that there are likely many truly affected individuals in these groups. The large number of individuals reporting diagnoses of all three diseases who have no record of the disease in the EHR database is particularly concerning. These groups similarly show elevated carrier frequencies relative to controls, suggesting that a substantial proportion of these self-reports are likely to be accurate. This highlights the degree of missingness in the EHRs in general and how much potential utility of a resource like *AoU* is being lost to the bottleneck of EHR-based phenotyping.

**Table 3.** Stratifying case counts and pLoF carrier frequencies by whether participants indicated disease history in Personal and Family Health History Survey.

|  | Indicated disease in survey? | Confident cases |  | Tentative cases: set 1 |  | Tentative cases: set 2 |  | Controls |  |
| --- | --- | --- | --- | --- | --- | --- | --- | --- | --- |
|  |  | Count | Freq. | Count | Freq. | Count | Freq. | Count | Freq. |
| **Ovarian cancer** | Yes | 136 | 13.2% | 55 | 10.9% | 81 | 11.1% | 150 | 3.3% |
|  | No | 232 | 7.3% | 120 | 6.0% | 432 | 3.2% | 95,192 | 0.5% |
| **Breast cancer** | Yes | 2,835 | 4.5% | 243 | 2.9% | 248 | 3.8% | 1,314 | 2.8% |
|  | No | 2,666 | 3.4% | 656 | 3.8% | 698 | 3.7% | 94,261 | 0.5% |
| **Colorectal cancer** | Yes | 71 | 1.4% | 301 | 3.2% | 349 | 2.9% | 485 | 0.6% |
|  | No | 138 | 2.2% | 595 | 2.7% | 1,295 | 1.6% | 157,317 | 0.1% |

Phenotype definition: = eMERGE = Rule of two = Single EHR

These results also highlight a useful role that self-reported survey data has the potential play in EHR-linked biobanks: countering the demographic biases associated with structural biases in EHRs. Ovarian cancer analysis in *AoU* provides an illustrative example. Based on our analysis, researchers seeking to maximize statistical power would be encouraged to adopt a higher-specificity case definition biased toward White women. Ideally, survey data would provide a supplementary route in conjunction with EHR-based evidence to assist in identifying cases in historically medically underserved populations who are more likely to be missed by the phenotyping approaches considered here. However, the set of individuals who responded to the optional Personal and Family Health History survey provided by *AoU* are disproportionately White (71% White, 10% Black) relative to the overall srWGS cohort (53% White, 21% Black), limiting its utility in this regard. Incorporating personal health history into the portion of the survey required by all participants could serve as a useful supplement to diversify case sets.

While the LoF-based phenotyping evaluation approach used here could be extended to include additional lower-penetrance risk alleles identified through common variant genome wide association studies (GWAS), there are diminishing returns in improved precision along with potential issues introduced by doing so. The framework provided here serves as a useful and simple heuristic for guiding case set decisions across a range of phenotypes in large EHR-linked biobanks like *AoU* via a misclassification estimator that is likely to be independent of downstream association testing. One risk in expanding the set of estimator alleles is the potential to break this independence assumption by estimating misclassification proportions using variants that are correlated with sites of interest. Additionally, since the overwhelming majority of GWAS participants are of European ancestry, expanding the variant set to top GWAS hits risks developing an estimator that is biased toward selecting the candidate case set with a greater proportion of EUR-origin individuals, further exacerbating the issued discussed above. A simple, high-penetrance estimator avoids these pitfalls while retaining most of the potential utility.

## Conclusions

The approach outlined here provides a useful first approximation for evaluating case ascertainment accuracy and produces recommendations for developing case sets in the *AoU* v7 cohort for EOC, breast cancer, and CRC. The choice of phenotyping strategy in large EHR-linked biobanks like *AoU* can have substantial implications for downstream statistical power, particularly for rare diseases. Additionally, requiring a greater depth of EHR-based evidence to define cases risks biasing case sets in favor of white individuals of EUR ancestry. Evidence suggests that self-reported survey responses indicating disease status may serve as a useful supplement to EHR-based evidence.

## Supporting information

Figure S1, Table S1, etc

## Data Availability

The data used in this study are available to registered researchers from the All of Us Research Program (https://allofus.nih.gov/). All analyses were performed on the All of Us researcher workbench. Scripts are available upon request.

## Acknowledgements

This work was supported by Tell Every Amazing Lady (T.E.A.L.) Medical Research Grant. We thank *All of Us* participants for their contributions. We also thank the *All of Us* Research Program for making available the participant data examined in this study.

## Data and code availability

The data used in this study are available to registered researchers from the *All of Us* Research Program (https://allofus.nih.gov/). All analyses were performed on the *All of Us* researcher workbench. Scripts are available upon request.

## References

1. Hersh, W. R. et al. Caveats for the Use of Operational Electronic Health Record Data in Comparative Effectiveness Research. Med. Care 51, S30–S37 (2013).

2. Getzen, E., Ungar, L., Mowery, D., Jiang, X. C Long, Q. Mining for equitable health: Assessing the impact of missing data in electronic health records. J. Biomed. Inform. 139, 104269 (2023).

3. Boyd, A. D. et al. Equity and bias in electronic health records data. Contemp. Clin. Trials 130, 107238 (2023).

4. Abramowitz, S. A. et al. Evaluating Performance and Agreement of Coronary Heart Disease Polygenic Risk Scores. JAMA (2024) doi:10.1001/jama.2024.23784.

5. Aschebrook-Kilfoy, B. et al. An Overview of Cancer in the First 315,000 All of Us Participants. PLOS ONE 17, e0272522 (2022).

6. Acosta, J. N. et al. Cardiovascular Health Disparities in Racial and Other Underrepresented Groups: Initial Results From the All of Us Research Program. J. Am. Heart Assoc. 10, e021724 (2021).

7. Shetty, N. S. et al. Titin truncating variants, cardiovascular risk factors and the risk of atrial fibrillation and heart failure. *Nat*. Cardiovasc. Res. 3, 899–906 (2024).

8. Bates, B. A. et al. Intersection of rare pathogenic variants from TCGA in the All of Us Research Program v6. Hum. Genet. Genomics Adv. 6, 100405 (2025).

9. Duan, R. et al. An Empirical Study for Impacts of Measurement Errors on EHR based Association Studies. AMIA Annu. Symp. Proc. AMIA Symp. 2016, 1764–1773 (2016).

10. ICD-10-CM official guidelines for coding and reporting FY 2023 -- UPDATED April 1, 2023 (October 1, 2022 - September 30, 2023). (2023).

11. Kirby, J. C. et al. PheKB: a catalog and workflow for creating electronic phenotype algorithms for transportability. J. Am. Med. Inform. Assoc. 23, 1046–1052 (2016).

12. Ye, Z. et al. Phenome-wide association studies (PheWASs) for functional variants. Eur. J. Hum. Genet. 23, 523–529 (2015).

13. Verma, A. & Ritchie, M. D. Current Scope and Challenges in Phenome-Wide Association Studies. Curr. Epidemiol. Rep. 4, 321–329 (2017).

14. Bastarache, L. Using Phecodes for Research with the Electronic Health Record: From PheWAS to PheRS. Annu. Rev. Biomed. Data Sci. 4, 1–19 (2021).

15. Yan, C. et al. Large language models facilitate the generation of electronic health record phenotyping algorithms. J. Am. Med. Inform. Assoc. 31, 1994–2001 (2024).

16. Wei, W.-Q. C Denny, J. C. Extracting research-quality phenotypes from electronic health records to support precision medicine. Genome Med. 7, 41 (2015).

17. KPWA/UW. Ovarian/Uterine Cancer (OvUtCa). (2017).

18. Shang, N., Hripcsak, G., Weng, C., Chung, W. K. & Crew, K. Breast Cancer. (2018).

19. Carrell, D. & Grafton, J. Colorectal Cancer (CRC). (2016).

20. Antoniou, A. et al. Average Risks of Breast and Ovarian Cancer Associated with BRCA1 or BRCA2 Mutations Detected in Case Series Unselected for Family History: A Combined Analysis of 22 Studies. Am. J. Hum. Genet. 72, 1117–1130 (2003).

21. Frank, T. S. et al. Clinical Characteristics of Individuals With Germline Mutations in *BRCA1* and *BRCA2* : Analysis of 10,000 Individuals. J. Clin. Oncol. 20, 1480–1490 (2002).

22. Phelan, C. M. et al. A low frequency of non-founder BRCA1 mutations in Ashkenazi Jewish breast-ovarian cancer families: ASHKENAZI JEWISH NON-FOUNDER BRCA1 MUTATIONS. Hum. Mutat. 20, 352–357 (2002).

23. McClain, M. R., Palomaki, G. E., Nathanson, K. L. & Haddow, J. E. Adjusting the estimated proportion of breast cancer cases associated with BRCA1 and BRCA2 mutations: Public health implications. Genet. Med. 7, 28–33 (2005).

24. Maxwell, K. N., Domchek, S. M., Nathanson, K. L. & Robson, M. E. Population Frequency of Germline *BRCA1/2* Mutations. J. Clin. Oncol. 34, 4183–4185 (2016).

25. Ramus, S. J. & Gayther, S. A. The Contribution of *BRCA1* and *BRCA2* to Ovarian Cancer. Mol. Oncol. 3, 138–150 (2009).

26. Hall, M. J., et al. *BRCA1* and *BRCA2* mutations in women of different ethnicities undergoing testing for hereditary breast-ovarian cancer. Cancer 115, 2222–2233 (2009).

27. Haffty, B. G. Racial differences in the incidence of BRCA1 and BRCA2 mutations in a cohort of early onset breast cancer patients: African American compared to white women. J. Med. Genet. 43, 133–137 (2005).

28. Lynch, H. et al. Review of the Lynch syndrome: history, molecular genetics, screening, differential diagnosis, and medicolegal ramifications. Clin. Genet. 76, 1–18 (2009).

29. Prevalence and penetrance of BRCA1 and BRCA2 mutations in a population-based series of breast cancer cases. Br. J. Cancer 83, 1301–1308 (2000).

30. Bonadona, V. Cancer Risks Associated With Germline Mutations in MLH1, MSH2, and MSH6 Genes in Lynch Syndrome. JAMA 305, 2304 (2011).

31. Edwards, B. J., Haynes, C., Levenstien, M. A., Finch, S. J. & Gordon, D. Power and sample size calculations in the presence of phenotype errors for case/control genetic association studies. BMC Genet. 6, 18 (2005).

32. Abu-Ghazaleh, N., Kaushik, V., Gorelik, A., Jenkins, M. & Macrae, F. Worldwide prevalence of Lynch syndrome in patients with colorectal cancer: Systematic review and meta-analysis. Genet. Med. 24, 971–985 (2022).

33. The All of Us Research Program Investigators. The “All of Us” Research Program. N. Engl. J. Med. 381, 668–676 (2019).

34. Kerminen, S. et al. Geographic Variation and Bias in the Polygenic Scores of Complex Diseases and Traits in Finland. Am. J. Hum. Genet. 104, 1169–1181 (2019).

35. Reisberg, S., Iljasenko, T., Läll, K., Fischer, K. & Vilo, J. Comparing distributions of polygenic risk scores of type 2 diabetes and coronary heart disease within different populations. PLOS ONE 12, e0179238 (2017).

36. Yland, J. J., Wesselink, A. K., Lash, T. L. & Fox, M. P. Misconceptions About the Direction of Bias From Nondifferential Misclassification. Am. J. Epidemiol. 191, 1485–1495 (2022).

37. Chyou, P.-H. Patterns of bias due to differential misclassification by case–control status in a case–control study. Eur. J. Epidemiol. 22, 7–17 (2007).

38. Mitra, S. K. On the Limiting Power Function of the Frequency Chi-Square Test. Ann. Math. Stat. 29, 1221–1233 (1958).

