## Supplementary material for "Measuring the accuracy of electronic health record (EHR)-based phenotyping in the *All of Us* Research Program to optimize statistical power for genetic association testing": Figure S1, Table S1, etc

### Supplemental methods

#### Diagnosis codes in *All of Us*

To harmonize record across health care systems, *AoU* utilizes the Observational Medical Outcomes Partnership (OMOP) Common Data Model. All medical records from any source vocabulary (e.g. ICD-9-CM, SNOMED-CT) are standardized by mapping codes onto the Standardized Nomenclature of Medicine (SNOMED) ontology. However, the phenotyping algorithms considered in this analysis rely primarily on ICD-9-CM or ICD-10-CM codes to identify cases. To account of for this, we extend the published eMERGE pipelines to also include any source code that maps onto the same standardized SNOMED code as one of the qualifying ICD-9-CM or ICD-10-CM code for the target disease. An example is shown in *fig. S1*. This approach effectively casts as wide of a net as possible for each relevant code within the *AoU* database. Additionally, since recordkeeping norms may vary across hospital systems, this ensures that those utilizing vocabularies other than ICD-9-CM or ICD-10-CM are not systematically excluded from the analysis. The additional codes included by this approach were manually reviewed to ensure reasonableness.

#### Calculating power with misclassified outcomes

A simple burden test using a logistic regression model to test association between binary loss of function status and the disease of interest is equivalent to a chi-square test of association. Mitra (1958) showed that misclassification can be accounted for by expressing the non-centrality parameter of the non-central chi-square distribution, $\lambda$, in terms of the probabilities of true disease status among cases and controls[38]. Letting $n_1$ and $n_0$ denote the numbers of cases and controls, respectively:

$$\lambda = n_1 n_0 \sum_{j=0}^{1} \frac{\left(p_{0j} - p_{1j}\right)^2}{n_1 p_{1j} + n_0 p_{0j}} \tag{2}$$

where $p_{1j}$ and $p_{0j}$ give the frequencies of allele *j* in cases and controls, respectively. For a loss of function burden test, $j \in \{0,1\}$.

Since the case groups consist of a mixture of unaffected and affected individuals, under the law of total probability, the frequency of allele *j* in the case and control groups can be expressed in terms of the probability an affected individual is classified as a control ($\theta$), the probability an unaffected individual is classified as a case ($\phi$), the disease prevalence (*K*), and the frequencies of allele *j* in cases and controls ($p_{Aj}$ and $p_{Uj}$, respectively):

$$p_{1j} = \frac{p_{Aj}(1-\theta)K + p_{Uj}\phi(1-K)}{(1-\theta)K + \phi(1-K)} \tag{3}$$

$$p_{0j} = \frac{p_{Aj}\theta K + p_{Uj}(1-\theta)(1-K)}{\theta K + (1-\phi)(1-K)} \tag{4}$$

Estimating these parameters allows us to compute downstream statistical power under the tentative case definitions. Under all criteria, we assume that $\theta = 0$ since a highly conservative non-cancer control definition is used in both analyses. Additionally, the large control set sizes for all three phenotypes make this assumption robust to violations at the scale we might expect for the diseases studied here. In the case where were the observed carrier frequency is greater in tentative cases than in confident cases, we cap the estimated PPV at 1 ($\widehat{PPV} = \min\{\widehat{n_A}/n_1, 1\}$). We then estimate $\phi$ as:

$$\hat{\phi} = \frac{n_1\left(1 - \widehat{PPV}\right)}{n_0 + n_1\left(1 - \widehat{PPV}\right)} \tag{5}$$

There are several reasonable options for estimating $p_{Aj}$. One approach would be to use approximations from prior population-based studies. Another would be to take the observed carrier frequency in the confident case set as an estimate by assuming perfect specificity under that case definition. We use the latter approach in this analysis, opting for an empirical estimate computed directly from the *AoU* cohort. This produces reasonable values in line with prior population-based estimates, so there would be little difference between these approaches. We use observed carrier frequencies in non-cancer controls to estimate $p_{Uj}$.

#### Supplemental figures

***Table S1 | Comparing proportions of non-Hispanic White (White) and Black individuals by case set***

| | **Confident cases** | | **Tentative cases: set 1** | | **Tentative cases: set 2** | | **Controls** | |
|---|---|---|---|---|---|---|---|---|
| | Black | White | Black | White | Black | White | Black | White |
| **Ovarian cancer** | 8% | 70% | 14% | 70% | 14% | 63% | 21% | 51% |
| **Breast cancer** | 11% | 71% | 12% | 63% | 13% | 63% | 19% | 52% |
| **Colorectal cancer** | 9% | 69% | 9% | 69% | 12% | 69% | 21% | 51% |

***Figure S1 | Applying ICD code-based phenotyping procedures in AoU requires harmonizing codes across ontologies and health care systems:*** *The AoU EHR database maps records from all source vocabularies onto the SNOMED ontology. The algorithmic EHR pipelines used here rely on ICD-9-CM and ICD-10-CM codes. To harmonize these, we apply these algorithms using their mapped equivalent SNOMED codes, followed by a manual review to assume no irrelevant diagnoses or conditions are contributing to the final case set*

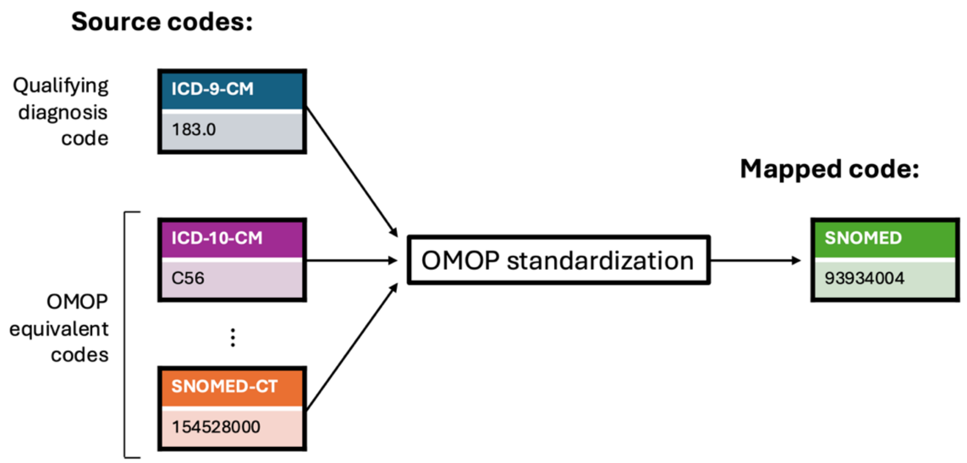

***Figure S2 | Histograms of age at diagnosis by disease and case set in AoU v7***

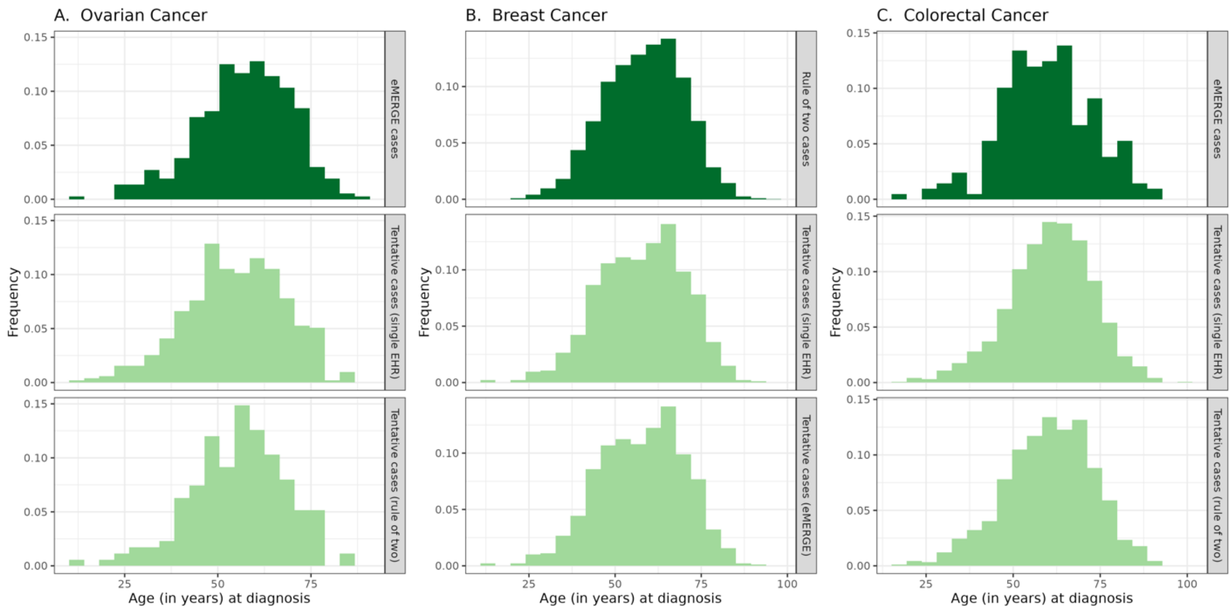

***Figure S3 | Counts of individuals in AoU v7 qualifying as cases under each combination of four possible phenotyping approaches.*** *These show mutually exclusive groups that can be combined to produce the counts in tables 2 and 3 of the main text.*

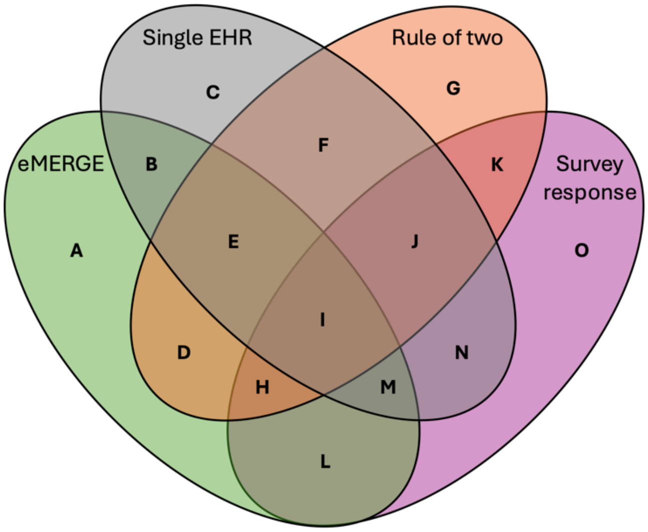


| | EOC | Breast | CRC |
|---|---|---|---|
| **A** | 0 | 0 | 0 |
| **B** | 0 | 656 | 25 |
| **C** | 312 | 42 | 700 |
| **D** | 0 | 0 | 0 |
| **E** | 232 | 2,666 | 113 |
| **F** | 120 | 0 | 595 |
| **G** | 0 | 0 | 0 |
| **H** | 0 | 0 | 0 |
| **I** | 136 | 2,835 | 64 |
| **J** | 55 | 0 | 301 |
| **K** | 0 | 0 | 0 |
| **L** | 0 | 0 | 0 |
| **M** | 0 | 243 | 7 |
| **N** | 26 | 5 | 48 |
| **O** | 150 | 1,314 | 485 |
